# *Plasmodium falciparum* genomic surveillance reveals a diversity of *kelch13* mutations in Zambia

**DOI:** 10.1101/2025.02.19.25322554

**Authors:** Andrés Aranda-Díaz, Sydney Mwanza, Takalani I. Makhanthisa, Sonja B. Lauterbach, Faith De Amaral, Mukosha Chisenga, Brighton Mangena, Isobel Routledge, Blaženka Letinić, Bertha Kasonde, Gershom Chongwe, Mulenga C. Mwenda, John M. Miller, Tricia Hibwato, Chirwa Jacob, Busiku Hamainza, Stephen Bwalya, Japhet Chiwaula, Japhet Matoba, Chadwick Sikaala, John Chimumbwa, Amy Wesolowski, Jennifer L. Smith, Jaishree Raman, Moonga Hawela

**Author notes:** Corresponding author. Address: C/ Rosselló, 132, 7th 2^nd^, 08036, Barcelona, Spain. Co-senior authors (Jennifer L. Smith, Jaishree Raman, Moonga Hawela).

## Abstract

Antimalarials are central to Zambia’s strategies for malaria control and elimination. Antimalarial drug resistance poses a significant threat to the effectiveness of artemisinin-based combination therapies and preventive strategies such as sulfadoxine-pyrimethamine chemoprevention in pregnant women. In this genomic surveillance study, dried blood spots and epidemiological data were collected from confirmed Plasmodium falciparum cases at 61 health facilities across all 10 Zambian provinces from March to July 2023. A total of 2,486 samples were genotyped using multiplexed amplicon sequencing to identify mutations in 12 genes associated with resistance to seven antimalarial drugs. Several mutations potentially associated with artemisinin partial resistance were identified, including the validated k13 P574L marker (0.66% adjusted national prevalence) and the candidate k13 P441L marker (1.39%). The distribution of mutations was heterogeneous, with many health facilities reporting resistance markers in more than 5% of infections, and in some instances, up to 46%. The mdr1 N86 genotype, associated with decreased lumefantrine susceptibility, was found in all samples. Very high levels of sulfadoxine-pyrimethamine resistance markers were observed, including dhps K540E (93.26%). The variable prevalence of resistance markers underscores the need for routine molecular surveillance to detect emergent resistance and guide malaria control strategies. These results also call for studies to understand the clinical implications of these mutations and ensure the continued efficacy of antimalarial interventions in Zambia.

## Introduction

Between 2000 and 2015, malaria mortality and morbidity reduced globally, with cases declining from 79 to 58 per 1,000 population at risk.^1^ This progress was driven by the widespread adoption of effective vector control interventions (e.g. long-lasting insecticide-treated nets, indoor residual spraying), improved case management strategies (e.g. prompt diagnosis with rapid diagnostic tests (RDTs) and effective treatment of malaria infections), and expanded access to preventative treatment for vulnerable populations (e.g. seasonal malaria chemoprevention, intermittent treatment for pregnant women). Despite these gains, malaria remains a critical public health concern, particularly in sub-Saharan Africa which accounts for approximately 95% of global cases and deaths.^1^ Progress has plateaued due to a multitude of operational and biological challenges.^1^ Among them, the emergence of antimalarial drug resistance has been identified as a critical driver of malaria transmission, posing a significant threat to global malaria control and elimination efforts.^2,3^

Globally, antimalarial drug resistance has emerged against all frontline antimalarials, including artemisinin-based combination therapies (ACTs), the malaria treatment currently recommended by the World Health Organization. Zambia, a country in southern Africa that has highly heterogeneous rates of malaria transmission, was one of the first countries in sub-Saharan Africa to adopt an ACT, artemether-lumefantrine, as the first-line treatment for uncomplicated malaria in response to widespread resistance to chloroquine.^4^ Artemisinin partial resistance (ART-R), defined by delayed parasite clearance following treatment, is widespread in Southeast Asia and has recently emerged in East, Central-East Africa, and the Horn of Africa.^5,6^ Resistance to ACT partner drugs appears to be limited in Africa, with recent data suggesting that artemether-lumefantrine and dihydroartemisinin-piperaquine, used in mass treatment campaigns, remain efficacious in Zambia.^7,8^ Although sulfadoxine-pyrimethamine (SP)-resistant parasites are prevalent in the country, SP remains effective for the prevention of malaria in pregnancy in Zambia.^7,9–16^

Therapeutic and chemoprevention efficacy studies are crucial for monitoring drug effectiveness but are labor-intensive and costly. Molecular surveillance for mutations associated with antimalarial drug resistance can complement these studies by cost-efficiently quantifying spatiotemporal trends in drug resistance marker prevalence, and identifying areas of concern that therapeutic efficacy studies could target. Certain mutations known as single nucleotide polymorphisms (SNPs) in the *kelch* 13 (*k13)* gene, result in amino acid changes in the propeller domain of the K13 protein which are associated with ART-R.^17,18^ Over the last decade, different *k13* SNPs associated with ART-R have emerged in the Horn of Africa, Central Africa, and East Africa.^19–24^ In contrast, chloroquine resistance is mediated primarily by a fixed set of mutations in the chloroquine resistance transporter (*crt*) gene and is modulated by mutations in the multidrug resistance 1 (*mdr1*) gene.^25^ Mutations in *crt* and *mdr1* genes are also associated with resistance to piperaquine, amodiaquine, and lumefantrine.^26,27^ An accumulation of specific SNPs in the dihydropteroate synthetase (*dhps*), and dihydrofolate reductase (*dhfr*) genes confer resistance to SP.^28^

Despite the high prevalence of *k13* SNPs and recent reports of delayed parasite clearance in the Horn of Africa, Central Africa, and East Africa, molecular surveillance and therapeutic efficacy studies in southern Africa are limited.^19–24^ In Zambia, up until 2018, molecular surveillance studies using either Sanger or molecular inversion probe deep sequencing have rarely detected any *k13* mutations.^11,12,29^ Conversely, the *crt* K76T chloroquine resistance marker was found at low levels, while markers of resistance to sulfadoxine-pyrimethamine were prevalent across the country.^9–13^ To provide a more comprehensive description of antimalarial drug resistance marker prevalence across southern Africa to help guide treatment policy and responses, the Genomics for Malaria in the Elimination 8 (GenE8) initiative implemented malaria molecular surveillance using targeted amplicon deep sequencing in 5 southern African countries (Angola, Eswatini, Namibia, South Africa, and Zambia). Here, we report on the prevalence of molecular markers associated with resistance to commonly used antimalarials in all 10 Zambian provinces, based on falciparum-positive samples collected between March and July 2023.

## Materials and Methods

### Study Site

Zambia is a landlocked country in southern Africa that neighbors the Democratic Republic of the Congo, Angola, Namibia, Botswana, Zimbabwe, Mozambique, Malawi, and Tanzania. Almost the entire population of 19 million (99%) is at risk of contracting malaria, with an average malaria prevalence of 29.3% among 2 to 10-year-olds.^7^ Malaria transmission is highly heterogenous displaying a north-to-south gradient. The northern regions, including Luapula province (63.3%), North-Western (47.4%), Western (47.4%), and Muchinga provinces (43.8%) had the highest prevalence in 2021, while Southern and Lusaka provinces in the south are considered to be in pre-elimination with a prevalence of 3.3% (Figure 1A).^7,30^ Malaria transmission is year-round but peaks between January and April. Most (98%) infections are due to *Plasmodium falciparum* mono infections, with co-infections of *P. falciparum* and *P. malariae*, *P. ovale,* and *P. vivax* occasionally reported.^7,31,32^ Common vectors are *Anopheles funestus*, *An. gambiae* s.s, and *An. arabiensis*.^7^ The health care system in Zambia includes 3,534 health facilities that serve as diagnostic and treatment centers for malaria. Malaria is diagnosed at point of care either by *P*. *falciparum* histidine-rich protein 2 (HRP2)-based RDTs or microscopy.^33^ All patients with uncomplicated malaria are treated with artemether-lumefantrine, while those with severe malaria are treated with intravenous artesunate.^7,33^. Dihydroartemisinin- piperaquine is reserved for mass drug administration in eliminating districts, while SP is used as an intermittent preventive treatment in pregnancy (IPTp).

**Figure 1.**
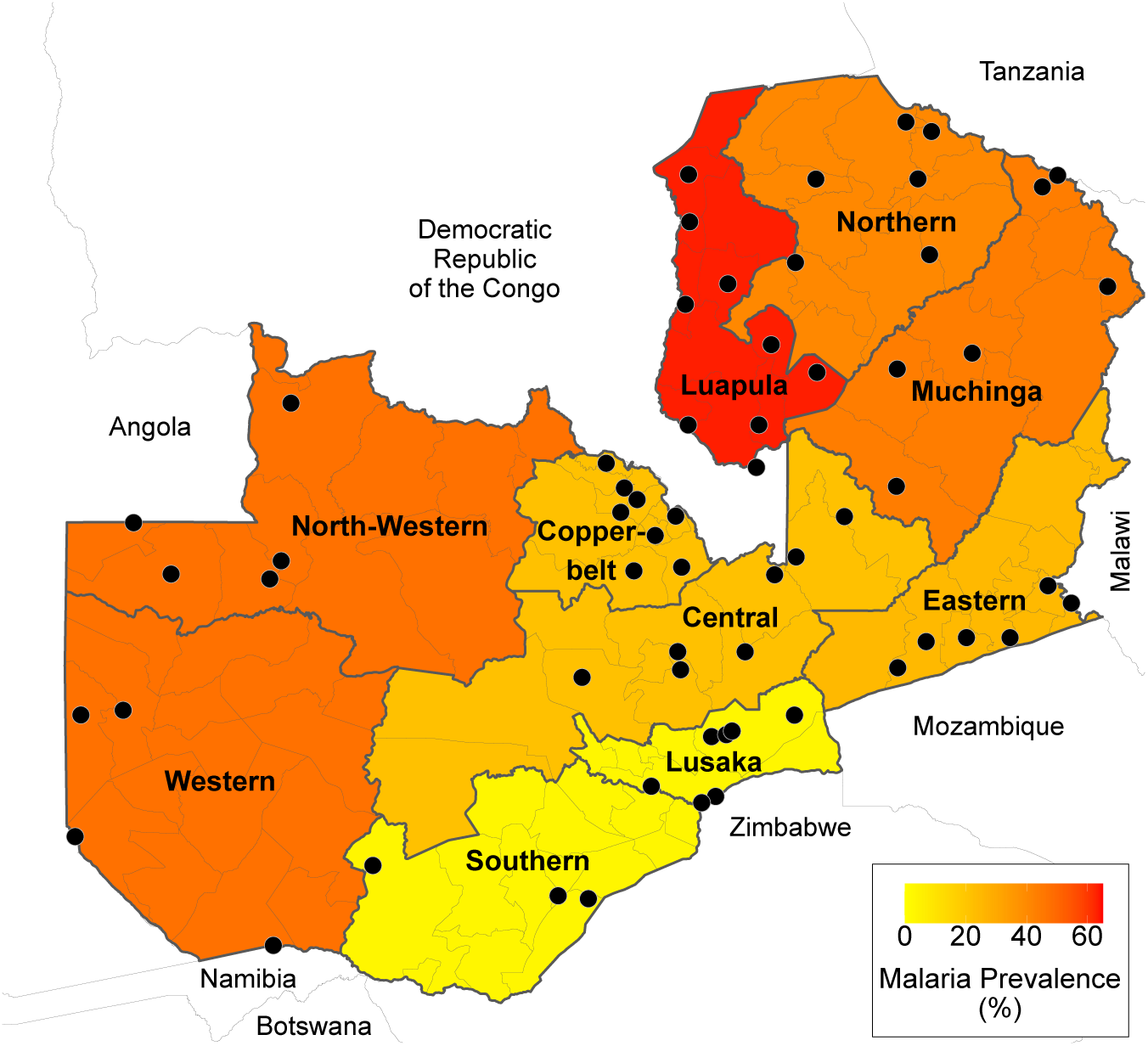
Study site. **A.** Map of Zambia with province-level malaria prevalence^30^ indicating 61 health facilities with successfully sequenced samples (black dots).

### Study Design, Sampling and Data Collection

This cross-sectional health facility-based study was conducted as part of the wider GenE8 project, and sequenced samples collected as part of the Regional GenE8 project (ReGenE8) and contemporaneous samples from other ongoing studies. The ReGenE8 project was nested within the national surveillance system and prioritized the representativeness of health facilities across geographies and pre-defined transmission strata. A minimum of 50 successfully sequenced samples from malaria-RDT-positive patients were targeted at each of the 20 selected health facilities, which are distributed across all 10 provinces. To achieve this, 200 malaria-RDT-positive patients were targeted for dry blood spot (DBS) and demographic data collection. The target sequencing sample size (n=1,000) was intended to: a) estimate a 5% prevalence of a given molecular marker with a precision of 1.65% and 95% confidence, and b) achieve 90% power to detect a prevalence exceeding a value of 2% if the true prevalence is 3.9% using a one-sided binomial test with ɑ=0.05 and a design effect of 1.5. An additional 2,000 samples from 33 health facilities under a country-specific GenE8 project award, as well as from 7 health facilities from a contemporaneous HRP2/3 surveillance project, were included to increase geographic representativeness. The achieved sample size of 2,013 sequenced infections across 61 health facilities provided a precision of 1.15% for a proportion of 5%, assuming a design effect of 1.5. The actual design effect was 14.60, and the intraclass correlation coefficient was 0.42.

Data collection was conducted from March to July during the 2023 malaria transmission season in 67 participating health facilities (Figure 1A). All symptomatic patients over the age of 2 years who tested positive for *falciparum* malaria with an HRP2-based RDT (SD Bioline Malaria Ag P.f or First Response Malaria Antigen P. falciparum HRP2 Card tests) were invited to participate in the study. Patients with severe falciparum malaria, severe malnutrition, or febrile conditions attributable to non-malarial diseases were excluded from the study.

Prior to enrollment in the study, written informed consent was obtained as described in the Ethical Considerations section. Enrolled participants were interviewed at the point of care using CommCare or KoBo collect software on encrypted tablets to capture information on demographics, clinical symptoms, travel history, and occupation. Finger prick DBS samples collected on filter paper strips (Cytiva Whatman 3MM Chr) were labeled with unique barcodes to allow for the linking of genomic and survey data. The DBS samples were air-dried, and stored individually in sealed bags with desiccant at -20°C until they were shipped to the regional sequencing hub at the National Institute for Communicable Diseases in South Africa or to the University of California, San Francisco.

### Sample Processing and Genomic Assays

DNA was extracted from at least 90 samples (one 6 mm punched DBS disk per sample) per health facility using the Chelex-Tween 20 method.^34^ Parasite densities were estimated using the varATS qPCR assay, with a minimum of 70 falciparum-positive samples from each health facility with a concentration of >100 parasite/μL randomly selected for downstream sequencing analysis.^35^ Where this target sample size could not be achieved using the 100 parasite/μL cut- off, samples with parasitemias as low as 10 parasite/μL were included. Targeted amplicon sequencing libraries were prepared using the MAD^4^HatTeR panel pools D1.1, R1.2, and R2.1, and sequenced in either a NextSeq 2000 or a NovaSeq X instrument using 150 paired-end reads.^36^ A custom bioinformatics pipeline was used to infer alleles across 241 targets.^36^ Sequences resulting from targets in the following genes are reported: *k13*, *crt*, *mdr1*, *mdr2*, *dhps*, *dhfr*, *coronin*, *arps10*, *pib7*, *fd*, *exo*, PF3D7_1322700. Microhaplotypes are reported for SNPs present within the same amplicon. Variations to the original protocol and bioinformatic pipeline specifications are described in Eloff, Aranda-Diaz et al, 2025.^37^

Further data cleaning, analysis, and visualization were conducted in R (version 4.3.1). Samples with less than 10,000 reads, targets with <50 reads, and SNPs with within-sample-allele-frequency (WSAF) <0.01 or ≤10 reads were excluded from the analysis. Consequently, SNPs observed in only one sample at WSAF <1, and SNPs observed in multiple samples but exclusively in a single sequencing run, were also excluded. Sequencing preparation batches with high contamination levels in either the negative or positive controls were excluded from all the analyses.

Non-falciparum targets were excluded from sequencing depth summaries. Otherwise, all resulting data were analyzed, including samples with partial coverage of SNPs, resulting in varying total sample numbers for each SNP assessed. Samples were considered successfully sequenced if a genotype was obtained for all 59 assessed *k13* SNPs. Multilocus haplotypes for polymorphisms across 5 independent targets in *dhfr*/*dhps* genes (*dhps* amino acids 436, 437, 540, 581, and 613; and *dhfr* 51, 59, 108 and 164) were inferred for each sample by combining all individual haplotypes. Samples with mixed genotypes were considered carriers of the highest resistance level haplotype they contained. Samples with mixed genotypes in > 2 amplicons were considered undetermined.

To evaluate patterns in the data and identify potential issues with sampling and library preparation, sample similarity was estimated using the root mean square error of WSAF for highly diverse targets (pool D1.1), followed by clustering by Density-Based Spatial Clustering of Applications with Noise (DBSCAN) with ε=0.2. Health facilities with possible DBS duplication were excluded from the analysis. These excluded facilities had multiple clusters of samples with clonality >2 and nearly identical WSAF across all loci, suggesting multiple DBSs were prepared using blood from a single patient.

### Statistical Analysis

Descriptive statistics for gender, age group (<5, 5-14, 15-24, 25-39, 40-59, >59 years old), occupation (including the self-identified category ‘minor’), overnight travel in the past 2 months, and country of residence were calculated. To enable statistical analysis, some variables were reclassified: farming, herding, and fishing occupations were combined into a single occupational category (agricultural workers); individuals aged 25 years or older were grouped into a single category; and travel reports (within the 2 months prior to sample collection) were categorized as either international or domestic. Differences between successfully sequenced samples and qPCR-positive samples, as well as among provinces, were assessed using Chi- squared tests.

The proportion of samples carrying a mutation (i.e. polyclonal samples with mixed genotypes counted as a carrier of the mutation) was calculated. The prevalence of infections carrying a genotype at the country and province level was estimated, accounting for clustered sampling in provincial strata, and applied survey weights to adjust for unequal sampling probabilities using the R ‘survey’ package (v.4.4.2). 95% confidence intervals were obtained with the ‘beta’ method.

Multivariate logistic regression was performed using a generalized linear mixed-effects model to assess associations between the presence of markers of interest (as a binary outcome) and key categorical variables, including age group, gender, travel history, and occupation. A random intercept for each health facility and fixed effects for all other variables were included. This analysis was restricted to variables with sufficient sample size for convergence.

The complexity of infection (COI) for each sample and population allele frequencies were estimated jointly for each district and province using moire v3.4.0.^38^ Microhaplotypes in all *P*. *falciparum* highly diverse targets (primer pool D1.1), as well as SNPs or microhaplotypes in drug resistance targets of interest, were used to infer COI and allele frequencies. The percentage of polyclonal infections was estimated as the mean of the individual probabilities of polyclonality.

Species-specific *ldh* targets in the panel were used to identify co-infections with non- *falciparum* species. The resulting sequences were used to remove unspecific amplification and to classify *P*. *ovale* reads into *P*. *ovale curtisi* or *P*. *ovale wallikeri*. Non-*falciparum* species were considered present if the combined reads from all *ldh* targets were > 100, with each species contributing > 1% of the total *ldh* targets reads.

### Ethical considerations

Ethics approval for the study was obtained from the Tropical Diseases Research Centre Research Ethics Committee (TDRC-REC) and permission to conduct the study was granted from the Zambia National Health Research Authority. Written informed consent was obtained from all patients aged 18 years and older, as well as from parents/guardians of children aged 2 to 11 years. For patients aged 12 to 18 years, verbal assent was obtained, in addition to parent/guardian consent. Original signed consent forms were securely stored, with copies provided to the participants or their parents/guardians. All personal identifiers were removed from survey data prior to storage on a password-protected cloud, while consent forms were kept in a restricted-access location.

## Results

DNA was extracted from 4,737 samples with sequencing libraries prepared for 3,084 (65.1%) samples. A total of 379 sequenced samples were excluded from further analysis: 274 samples from 5 health facilities suspected of DBS duplication, along with 105 samples from other health facilities, including all samples from one facility, due to potential cross-contamination. At least one SNP genotype was obtained for 91.9% (2,486/2,705) of the samples across 61 health facilities, with 2,013 (74.4%) successfully sequenced (with genotypes for all 59 *k13* SNPs).

Samples from all 10 provinces were successfully sequenced, though sequencing success rates varied across provinces (Table 1). Luapula province had the highest number of successfully sequenced samples, while Western province had the lowest. The median per-sample sequencing depth was 719,499 reads (IQR: 198,595-1,509,018), with a median per-target depth of 1,257 reads (IQR: 183-4,566). A median of 221 out of 238 targets (IQR: 186- 225) had > 100 reads. SNPs with the lowest genotyping success rates included *crt* 145, *crt* 218/220, and *mdr1* 86 (genotyped in 2,021, 1,015, and 2,023 samples, respectively). Nearly three-quarters of the samples analyzed (70.6%) were polyclonal, with a population mean COI of 3.05 (95% CI: 3.02-3.09, N=2,446). Mixed genotypes in at least one drug resistance marker were detected in 52% of samples. Non-falciparum species, including *P*. *malariae*, *P*. *ovale curtisi*, *P*. *ovale wallikeri*, and *P*. *vivax*, were rare, collectively observed in only 3.8% of samples (Table S1).

**Table 1.** Demographics of participants with successfully sequenced samples.

| Characteristic | Overall | Western | North-Western | Southern | Copper-belt | Lusaka | Central | Luapula | Northern | Muchinga | Eastern |
| --- | --- | --- | --- | --- | --- | --- | --- | --- | --- | --- | --- |
| **N** | 2013 | 151 | 210 | 167 | 249 | 183 | 240 | 299 | 176 | 166 | 172 |
| **Health facilities** | 61 | 4 | 5 | 5 | 8 | 5 | 7 | 9 | 6 | 6 | 6 |
| **Age** |  |  |  |  |  |  |  |  |  |  |  |
| **< 5 years** | 217 (11%) | 18 (12%) | 15 (7.1%) | 12 (7.2%) | 32 (13%) | 13 (7.1%) | 14 (5.8%) | 38 (13%) | 24 (14%) | 30 (18%) | 21 (12%) |
| **5 - 14 years** | 777 (39%) | 64 (42%) | 97 (46%) | 83 (50%) | 86 (35%) | 81 (44%) | 83 (35%) | 106 (36%) | 65 (37%) | 64 (39%) | 48 (28%) |
| **15 - 24 years** | 574 (29%) | 46 (30%) | 65 (31%) | 46 (28%) | 69 (28%) | 55 (30%) | 79 (33%) | 77 (26%) | 50 (28%) | 40 (24%) | 47 (27%) |
| **≥25 years** | 442 (22%) | 23 (15%) | 33 (16%) | 26 (16%) | 62 (25%) | 34 (19%) | 64 (27%) | 77 (26%) | 37 (21%) | 30 (18%) | 56 (33%) |
| **Unknown** | 3 | 0 | 0 | 0 | 0 | 0 | 0 | 1 | 0 | 2 | 0 |
| **Gender** |  |  |  |  |  |  |  |  |  |  |  |
| **Female** | 1,018 (51%) | 76 (50%) | 144 (69%) | 81 (49%) | 117 (47%) | 84 (46%) | 122 (51%) | 153 (51%) | 89 (51%) | 73 (44%) | 79 (46%) |
| **Male** | 994 (49%) | 75 (50%) | 66 (31%) | 86 (51%) | 132 (53%) | 99 (54%) | 118 (49%) | 145 (49%) | 87 (49%) | 93 (56%) | 93 (54%) |
| **Unknown** | 1 | 0 | 0 | 0 | 0 | 0 | 0 | 1 | 0 | 0 | 0 |
| **Occupation** |  |  |  |  |  |  |  |  |  |  |  |
| **Agricultural** | 345 (19%) | 17 (11%) | 10 (7.6%) | 24 (14%) | 40 (16%) | 32 (17%) | 54 (23%) | 79 (36%) | 33 (19%) | 25 (21%) | 31 (18%) |
| **Minor** | 776 (43%) | 63 (42%) | 81 (62%) | 69 (41%) | 115 (46%) | 90 (49%) | 79 (33%) | 78 (36%) | 85 (48%) | 61 (51%) | 55 (32%) |
| **Other** | 147 (8.1%) | 24 (16%) | 7 (5.3%) | 6 (3.6%) | 33 (13%) | 3 (1.6%) | 10 (4.2%) | 15 (6.9%) | 14 (8.0%) | 15 (13%) | 20 (12%) |
| **Student** | 241 (13%) | 26 (17%) | 8 (6.1%) | 32 (19%) | 34 (14%) | 12 (6.6%) | 35 (15%) | 36 (17%) | 20 (11%) | 15 (13%) | 23 (13%) |
| **Unemployed** | 296 (16%) | 21 (14%) | 25 (19%) | 36 (22%) | 27 (11%) | 46 (25%) | 62 (26%) | 9 (4.1%) | 24 (14%) | 3 (2.5%) | 43 (25%) |
| **Unknown** | 208 | 0 | 79 | 0 | 0 | 0 | 0 | 82 | 0 | 47 | 0 |
| **Country of residence** |  |  |  |  |  |  |  |  |  |  |  |
| **Zambia** | 1,782 (99%) | 151 (100%) | 122 (93%) | 167 (100%) | 249 (100%) | 183 (100%) | 239 (100%) | 215 (99%) | 176 (100%) | 119 (100%) | 161 (94%) |
| **Other** | 23 (1.3%) | 0 (0%) | 9 (6.9%) | 0 (0%) | 0 (0%) | 0 (0%) | 1 (0.4%) | 2 (0.9%) | 0 (0%) | 0 (0%) | 11 (6.4%) |
| **Unknown** | 208 | 0 | 79 | 0 | 0 | 0 | 0 | 82 | 0 | 47 | 0 |
| **Reported travel (past 2 months)** |  |  |  |  |  |  |  |  |  |  |  |
| **No Travel** | 1,872 (96%) | 148 (99%) | 208 (100%) | 158 (97%) | 203 (94%) | 179 (99%) | 226 (97%) | 251 (88%) | 169 (97%) | 165 (100%) | 165 (96%) |
| **Domestic** | 61 (3.1%) | 1 (0.7%) | 0 (0%) | 5 (3.1%) | 7 (3.3%) | 1 (0.6%) | 7 (3.0%) | 32 (11%) | 2 (1.1%) | 0 (0%) | 6 (3.5%) |
| **International** | 1 (<0.1%) | 0 (0%) | 0 (0%) | 0 (0%) | 0 (0%) | 0 (0%) | 0 (0%) | 0 (0%) | 0 (0%) | 0 (0%) | 1 (0.6%) |
| **Unspecified** | 10 (0.5%) | 0 (0%) | 0 (0%) | 0 (0%) | 5 (2.3%) | 0 (0%) | 0 (0%) | 1 (0.4%) | 4 (2.3%) | 0 (0%) | 0 (0%) |
| **Unknown** | 69 | 2 | 2 | 4 | 34 | 3 | 7 | 15 | 1 | 1 | 0 |

A substantial proportion of the participants self-identified as minors (43%), while the most commonly reported occupations among adults were agricultural work (19%) and unemployment (16%). No significant differences in gender, travel history, or country of residence were observed between individuals with qPCR-positive samples and those successfully sequenced (Table 1, Table S2). However, non-Zambian residents, participants over 25 years old, and participants from Southern and Eastern provinces were underrepresented among the successfully sequenced samples. Most sequenced samples came from children and young adults (39% ages 5-14, and 29% ages 15-24) of either gender. Only a small proportion of participants were non-Zambian residents (1.3%) or reported any travel history (3.7%), with just one case of international travel recorded.

### Markers of artemisinin partial resistance

Non-synonymous mutations in 12 amino acids within the K13 propeller domain were detected in 225 samples (9.1% of all samples sequenced, Table 2). The candidate marker P441L, the validated marker P574L, and R539I, A578S, R622T, and P667A/S mutations were found in >0.5% of the samples sequenced. The validated marker C469Y was present in one sample from North-Western province, while 2 samples, one in Western province and one in Southern province carried the A675V marker.

**Table 2.** Summary of evaluated variants. Artemisinin partial resistance (ART-R) markers are categorized as validated or candidate following the current World Health Organization classification (<u>WHO 2025</u>). Variants in other genes can be found in Table S7.

| Gene | Observed in >0.5% of the samples | Observed in ≤0.5% of the samples |
| --- | --- | --- |
| ***k13*** | <u>Validated ART-R markers</u><br>P574L<br><u>Candidate ART-R markers</u><br>P441L<br><u>ART-R evidence lacking</u><br>C469C, A578S, R622T, P667A, P667S | <u>Validated ART-R markers</u><br>C469Y, A675V<br><u>ART-R evidence lacking</u><br>K479I, S485N, A504A, N537S, R539I, R575R, E605E, Q613E, Q613Q, G690G |
| ***crt*** |  | F48L, C72Y, M74I, N75E, K76T, A220S |
| ***dhfr*** | N51I, C59R, S108N, I164L |  |
| ***dhps*** | S436A, A437G, K540E, A581G | S436F, A613S |
| ***mdr1*** | F184Y, D1246Y | N86Y, T371T |

The candidate marker P441L was detected in 71 samples (3.1% of all samples sequenced) from 10 health facilities spanning 9 districts and 5 provinces (Figure 2A, Table S3- 7), with an estimated national prevalence of 1.39% (CI 0.45-3.25%, Figure 2B). The mutation was most prevalent in Central (5.32%, CI 0.03-33.8%) and Western provinces (3.69%, CI 0- 32.67%), but less than 2.2% in Southern, Lusaka, and North-Western provinces. Students (OR: 5.41, CI 1.37-21.3, with respect to minors) and those who reported overnight domestic travel in the past 2 months were more likely to carry P441L (OR: 10.1, CI 1.58-64.5, Table S8). In the single health facility in Chibombo district of Central province, nearly half (47%; 46/98) of the analyzed samples carried the P441L mutation, with an allele frequency of 30.5% (CI 23.7- 38.3%, Figure 2A, S1). This health facility also recorded the highest proportion of unemployed participants (65%), leading to an overrepresentation of the unemployed among P441L carriers.

**Figure 2.**
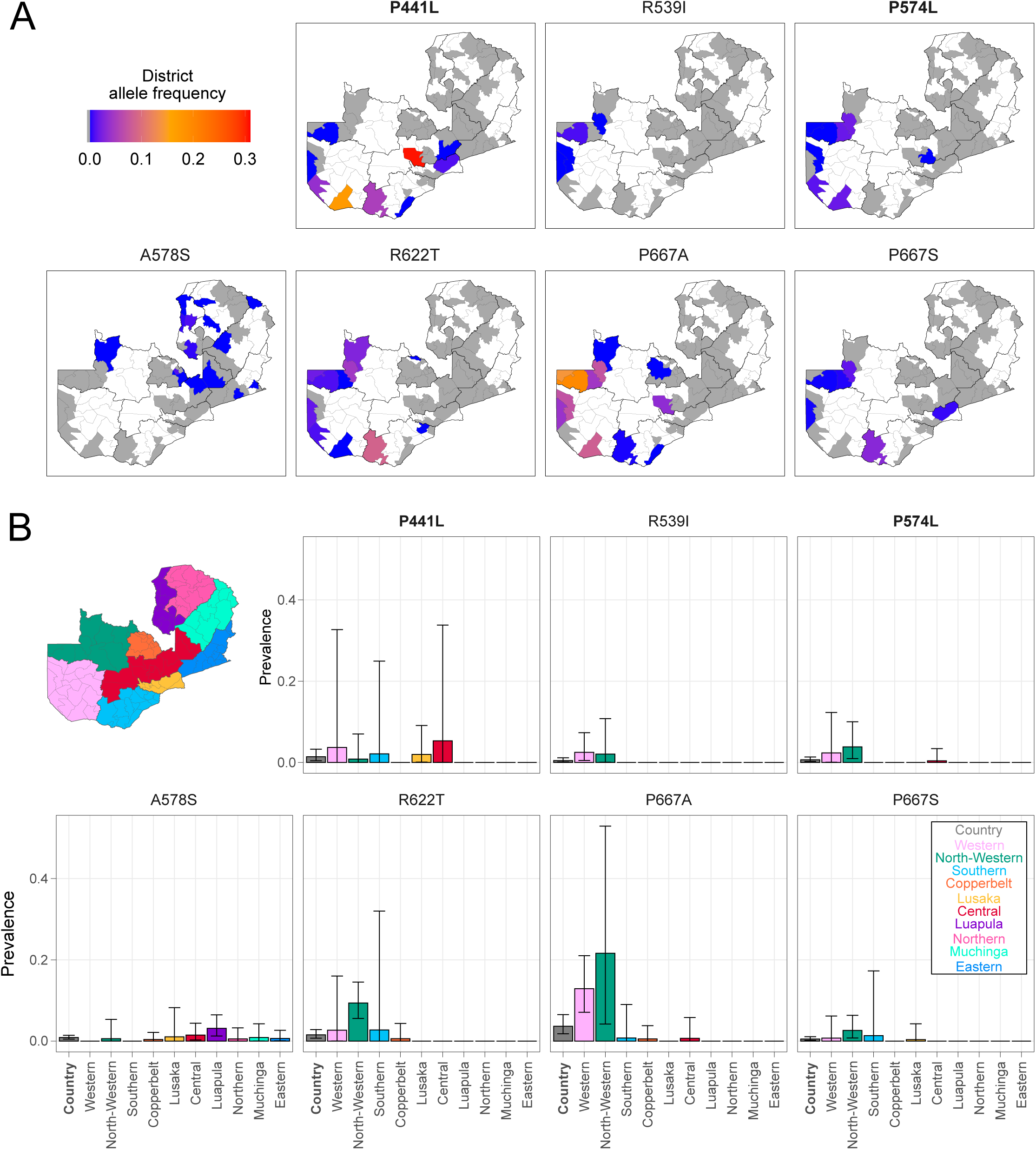
Allele frequency and prevalence of mutations in *k13*. **A**. District-level estimated allele frequencies for amino acids within *k13*. **B**. National- and province-level estimated prevalence of infections carrying a mutation. Error bars indicate 95% confidence interval. Candidate (P441L) and validated (P574L) markers are indicated in bold. Only SNPs with variants in >0.5% of the samples are shown.

The P574L marker was found in 16 samples (0.6% of the total) across 3 provinces (Western, North-Western, and Central provinces), with a national prevalence of 0.66% (CI 0.27- 1.34%). North Western and Western provinces had the highest proportions of P574L carriers and accounted for the majority of the observed R622T, R539I, P667A, and P667S mutations (Table S3). The P667A mutation reached a prevalence of 21.63% (CI 4.2-52.91%) in North- Western province, 12.91% (CI 7.09-21.01%) in Western province, and an estimated prevalence of 3.65% (CI 1.80-6.53%) nationally. The 667A allele reached frequencies of 19.5% (CI 10.6- 31.6%) and 13.8% (CI: 8.5-20.9%) in the neighboring districts of Zambezi and Chavuma in

North-Western province. National prevalences of the R622T and P667S mutations were lower (1.54%, CI 0.73-2.85%, and 0.51%, CI 0.19-1.09%, respectively). The A578S mutation was predominantly found in the eastern and northern provinces at a low national prevalence of 0.88% (CI 0.50-1.43%). Allele frequencies mirrored prevalence patterns across these markers (Table S4). There was no significant association between carriage of any mutation (validated or candidate markers, or mutations in the same amino acid as a validated or candidate marker) and demographic variables, including age, gender, or occupation (Table S8).

Other polymorphisms previously described as a genetic background in the emergence of *k13* mutations in South East Asia, including ferrodoxin (*fd*) D193Y, apicoplast ribosomal protein S10 (*arps10*) V127M, multidrug resistance protein 2 (*mdr2*) T484I and *crt* I356T were not detected in any samples (Table S7).^39^ Only one sample carried the phosphoinositide-binding protein 7 (*pib7*) gene mutation C1484F. Similarly, *coronin* mutations G50E, R100K and E107V associated with lower artemisinin resistance were absent in the data set.^40^

### Markers of SP resistance

Mutations associated with SP resistance were prevalent throughout the country (Figures 3 and S2, Tables S3-6). The SNPs within the triple *dhfr* mutant haplotype associated with pyrimethamine resistance, *dhfr* N51I, C59R and S108N, all had an estimated national prevalence above 90%. Southern province had the lowest *dhfr* N51I (81.59%, CI 64.42-92.82%) and *dhfr* C59R (76.97%, CI 50.79-93.41%) SNP prevalence. The *dhfr* S108N SNP was present in all samples, although 3 (0.86%) samples from Luapula province had a mixed genotype. Only two samples in Muchinga province carried *dhfr* I164L, a mutation that, when in a quadruple haplotype with the triple mutants, confers higher pyrimethamine resistance. Of the 2,379 samples with genotypes available in all 4 *dhfr* SNPs, 0.13% were undetermined (Figure S2). Of the 2,376 unambiguous samples, the triple mutant was found in 96.89%, the quadruple mutant in 0.08% (Figure 3C).

**Figure 3.**
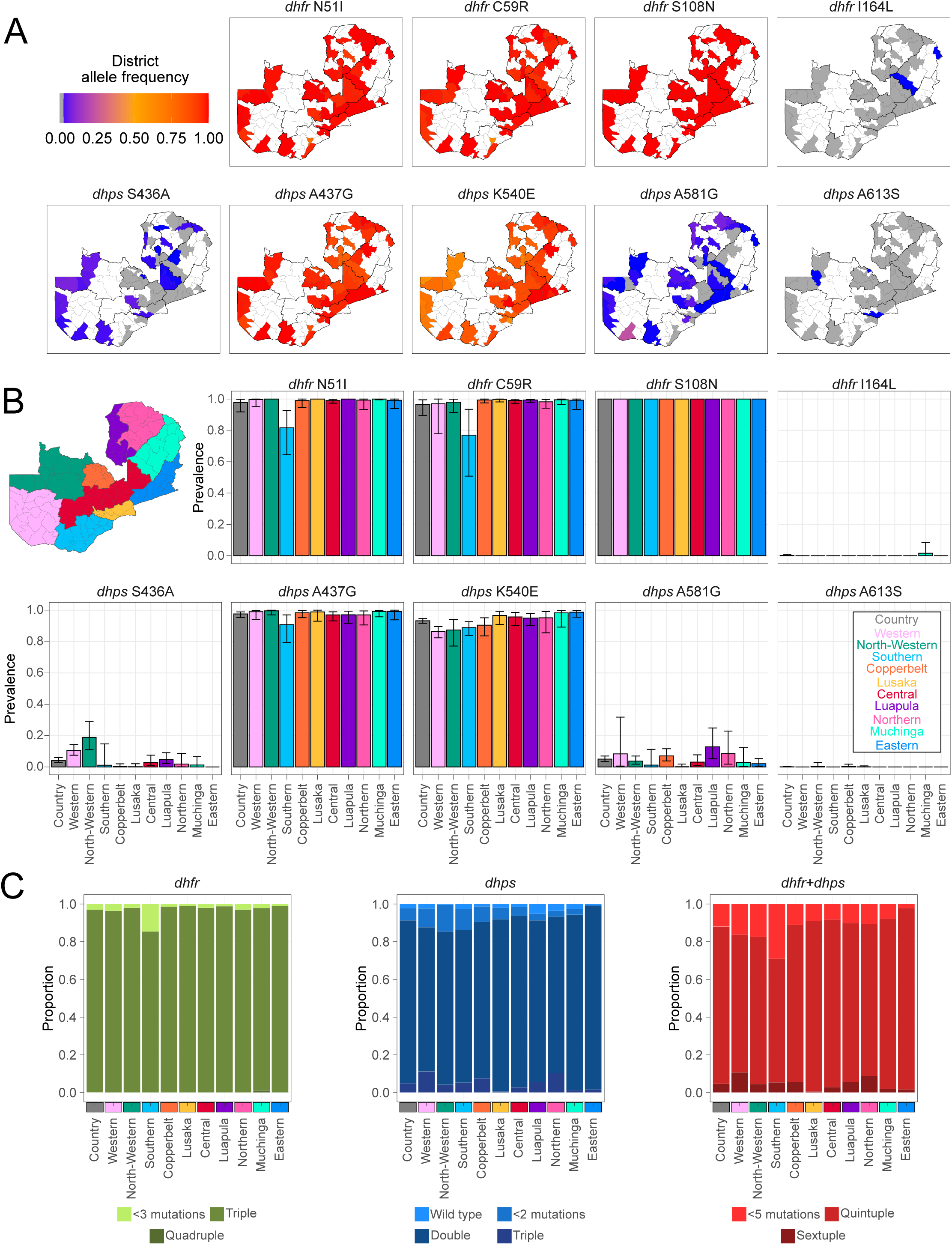
Allele frequency and prevalence of mutations in *dhfr* and *dhps*. **A**. District-level estimated allele frequencies for amino acids within *dhfr* and *dhps*. **B**. National- and province-level estimated prevalence of infections carrying a mutation. Error bars indicate 95% confidence interval. Only SNPs with variants in >0.5% of the samples are shown. **C**. Proportion of samples carrying *dhfr*, *dhps*, or *dhfr*/*dhps* haplotypes. Each sample was assigned the haplotype corresponding to the highest level of resistance detected. Polyclonal samples with mixed genotypes in multiple targets were undetermined and excluded (see Figure S2 for plots including these samples).

Of the *dhps* double mutant haplotype SNPs associated with sulfadoxine resistance - *dhps* S436A/F or A437G, and K540E - the *dhps* A437G and *dhps* K540E mutations had a national prevalence of 97.62% (CI 95.37-98.96%) and 93.26% (CI 91.59-94.69%), respectively. The *dhps* S436A mutant was found at 4.17% (CI 2.80-5.96%) nationally, while S436F was very rare (0.06%, CI 0-0.28%). The prevalence of the *dhps* A437G mutation was lowest in Southern province (90.78%, CI 79.42-97.04%), while *dhps* S436A was most prevalent in North-Western (18.87%, CI 11.02-29.12%) and Western provinces 10.52%, CI 7.48-14.27%). The *dhps* S436F mutation was only detected in Copperbelt and Lusaka provinces. The *dhps* K540E mutation had the lowest prevalence (<90%) in Western, North-Western, and Southern provinces. Two additional *dhps* mutations (A581G or A613S) associated with higher sulfadoxine resistance when part of a triple mutant haplotype, were found. The A581G mutation was found at 4.99% (CI 3.44-6.98%) nationally, with the highest prevalence in Luapula province (12.87%, CI 5.55- 24.2%) and a single health facility in Shesheke district, Western province, where it reached a proportion of 41.38% (12/29) and allele frequency of 22.1% (CI 11.8-35.6%). The A613S mutation was relatively rare, found in North-Western, Lusaka, and Copperbelt provinces at prevalences below 0.5%. Of the 2,257 samples with genotypes available in all 5 SNPs, 10.54% were undetermined (Figure S2). Of the 2,019 unambiguous samples, the double mutant haplotype was found in 86.38% and the triple mutant in 4.90% (Figure 3C). Reconstructing the full *dhps*/*dhfr* haplotype for 1,955 unambiguous samples found the quintuple mutant haplotype (*dhfr* triple and *dhps* double mutant haplotypes) in 83.43% and the sextuple mutant haplotype (*dhfr* quadruple and *dhps* double, or *dhfr* triple and *dhps* triple mutant haplotypes) in 4.55%.

### Markers of resistance to other antimalarial drugs

The *mdr1* N86 ‘wild type’ genotype, potentially selected for by lumefantrine use, was found in all samples.^41^ The *mdr1* N86Y mutation, associated with resistance to amodiaquine and chloroquine, was detected in only 4 samples. The *mdr1* Y184F and *mdr2* I492V mutations had prevalences of 50.08% (CI 42.63-57.53%) and 49.43% (CI 45.49-53.37%), respectively. Lastly, the chloroquine-resistant *crt* 72-76 CVIET microhaplotype has a relatively low national prevalence of 0.18% (CI 0.04-0.50%).

## Discussion

In this study, we investigated the prevalence of antimalarial drug resistance markers across all 10 provinces of Zambia using multiplexed amplicon deep sequencing. Mutations in the *k13* gene, which may impact artemisinin efficacy, were identified in six provinces. Validated markers such as C469Y, P574L, and A675V, along with the candidate marker P441L, were detected at low national prevalences (<3%) but showed notable geographic variation, with higher frequencies in specific health facilities and districts. Additionally, other *k13* mutations, including R622T and P667A/S, were observed. The *mdr1* N86 wild-type genotype, associated with lumefantrine tolerance, was present in all samples, while markers for SP resistance were widespread. Low-level co-infections with *P*. *malariae*, *P*. *ovale* curtisi, *P*. *ovale* wallikeri, and *P*. *vivax* were also detected.

Concerns around the emergence of partial artemisinin resistance in Africa have been driven by the rapid emergence of a diverse array of *k*13 mutations, including A675V, C469Y, P574L, and P441L, in the past decade in East Africa and the Horn of Africa.^19–23,42,43^ The *k13* A675V and C469Y markers, found in 2 and 1 Zambian samples respectively, have increased in prevalence in Uganda since 2016.^20^ In contrast, the validated marker P574L, rarely reported on the continent, was observed in 16 samples across 3 provinces in this study^44,45^ The candidate marker P441L, identified in 71 samples, has previously been reported in a few samples in Zambia’s North-Western province and the Democratic Republic of the Congo, and has recently increased in western Uganda.^20,29,46^ At the time of writing, a new study reported the emergence of P441L in Zambia’s Choma District, Southern province, with its prevalence rising from none in 2018 to 7.2% in 2023, consistent with our findings.^47^ A high proportion of P441L carriers has also been reported within the GenE8 project in Namibia’s Zambezi region, bordering Southern and Western provinces.^37^ Notably, the clinical and phenotypical effects of the P574L and P441L mutations on African parasites have not yet been evaluated.^2^ The P441L mutation, in particular, lacks robust clinical and laboratory evidence linking it to delayed clearance but a recent study in Myanmar observed an increase following mass drug administration with dihydroartemisinin- piperaquine.^48^ Other candidate or validated markers of ART-R that have been observed in the continent, such as R561H, C469F, and R622I, were not found in this study.^19,21–23,43^ However, we did find a high prevalence of R622T and P667A mutations, which occur in the same amino acids as the validated marker R622I and the mutations P667T/S, which have limited evidence of delayed clearance.^22,49,50^ The A578S mutation, which is widespread in Africa but does not impact artemisinin susceptibility, was observed at low levels and generally in areas where other *k13* mutations were not observed.^51^ The effect of other *k*13 mutations identified in this study potentially associated with delayed clearance also remains understudied.^29^

The study showed clear geographic trends in *k*13 mutations, with all mutations of interest observed in provinces in the western half of the country. The highest proportion of the P441L mutation was observed in Chibombo district, Central province, an intermediate transmission setting. ART-R has generally emerged in low transmission settings or areas experiencing a surge in transmission, where exposure of non-immune individuals can drive drug pressure due to treatment-seeking behavior and increase the likelihood of not clearing infections with resistant parasites.^20^ Furthermore, areas with low transmission are typically associated with lower intra-host genetic diversity, which would reduce within-host parasite competition and increase the likelihood of resistant strains persisting.^52,53^ This study also identified the P441L and P574L markers, as well as R622T and P667A/S, in intermediate transmission settings in Zambia, including a health facility near a Namibian facility where similar results were recently reported.^37^ These settings also exhibited higher levels of mixed polyclonal infections, consistent with the expected higher parasite genetic diversity in areas of intermediate malaria transmission. The interplay of factors - host immunity, pharmacokinetics, parasite physiology, epidemiology, and treatment-related non-intrinsic factors - in driving the selection and spread of these mutations remains to be fully understood.

The presence of ART-R does not necessarily result in ACT failure, as the partner drug can clear remaining parasites from the bloodstream. However, ART-R can compromise the efficacy of monotherapies used in severe malaria treatment and increase selection pressure for resistance to partner drugs. Currently, there is no strong evidence of full resistance to lumefantrine or pyronaridine, though resistance to other partner drugs such as amodiaquine, mefloquine, and piperaquine has been documented in South East Asia.^2^ In contrast, parasites in Africa generally lack the genotypes associated with resistance and have demonstrated continued high susceptibility in ex vivo studies.^54,55^ Decreased susceptibility to lumefantrine is associated with certain polymorphisms, including *crt* K76 and *mrd1* N86, both of which were observed in nearly all samples in this study.^27,54^ This likely reflects Zambia’s widespread use of artemether-lumefantrine for treatment of uncomplicated malaria. While modest changes in lumefantrine susceptibility have been reported in East Africa, they are unlikely to affect the efficacy of ACTs.^54,55^ No SNPs associated with resistance to amodiaquine or piperaquine were detected in this study. However, we did not assess copy number variations in *mdr1* and *plasmepsin* II/III, which confer resistance to mefloquine and piperaquine.^56,57^

Mutations associated with SP resistance were highly prevalent in this study. Although the full *dhps*/*dhfr* haplotype could not be determined in all samples due to sequencing depth variability, the quintuple mutant haplotype - a strong predictor of SP treatment failure - was detected in a majority of the samples. Its hallmark SNP, *dhps* K540E, was found at a prevalence exceeding 90%.^58^ While SP is not used as a partner drug for malaria treatment in Zambia, it is widely administered as IPTp, with approximately 90% of pregnant women receiving at least one dose.^30^ Notably, Southern province, which reported the lowest IPTp coverage (68.4%), also had the lowest prevalence of *dhfr* triple mutant haplotypes, largely due to a very low proportion of mutants in Pemba district.^30^ These findings suggest that SP is unlikely to be efficacious for malaria treatment nationwide and should not be considered as a partner drug in ACT regimens.

Despite the high prevalence of molecular resistance, SP remains effective for IPTp in Zambia, as multiple doses improve birth outcomes and clear infections even in settings with a high prevalence of the quintuple mutant haplotype.^14–16^ However, the effects of sextuple mutant haplotypes SNPs - including *dhps* A581G, *dhps* A613S, and *dhfr* I164L - on SP’s chemopreventive efficacy remain poorly understood. These SNPs were observed at an overall prevalence of approximately 5% but reached higher levels in specific districts within Luapula and Western provinces. Given SP’s importance in improving maternal health outcomes and its role in reducing selective pressure on ACT components, the efficacy of SP for IPTp should be closely monitored.

Parasite migration has played a critical role in the emergence and spread of antimalarial drug resistance in Africa and globally. For instance, chloroquine resistance first appeared in Southeast Asia and South America before rapidly spreading worldwide.^59^ Similarly, SP-resistant haplotypes originated in Southeast Asia and later migrated to Africa.^60^ In contrast, some mutations associated with ART-R have arisen independently in both Southeast Asia and Africa.^24,61^ The dynamics and drivers of ART-R spread in Africa remain poorly understood.

Specific ART-R molecular markers have been observed across borders in East Africa and the Horn of Africa, with sporadic reports elsewhere.^18–23,42,43,62^ Provinces in Zambia reporting *k13* mutations border Angola, Botswana, Mozambique, Namibia, and Zimbabwe, highlighting the potential role of parasite migration. Non-Zambian residents were captured in this study only in North-Western and Eastern provinces, but their numbers were insufficient for robust statistical analysis. The observed association between the *k13* P441L mutation and reported travel, which was almost exclusively domestic, is based on a limited number of cases and should be interpreted with caution. Previous research from Namibia’s Zambezi region, bordering Zambia’s Southern and Western provinces, underscored the role of human migration and parasite migration on malaria epidemiology in Namibia.^63,64^ Our findings on the spread of *k13* mutations in southwestern Zambia and northern Namibia align with these findings.^37^ Understanding the role of domestic and international human mobility in malaria transmission in Zambia is under-researched but critical for developing targeted strategies to limit the spread of resistance genotypes. On a larger scale, the relatedness of parasites carrying *k13* mutations in East Africa, the Horn of Africa, and Southeast Asia, to those found in this study remains unknown.

This study had several limitations. Samples were collected over part of a single season, limiting our ability to identify temporal trends. The sampling was designed to estimate country-level prevalence with broad geographical representation but was not powered to detect fine-scale geographical patterns. While most health facilities were randomly selected within provinces, some sentinel sites were intentionally added, potentially violating statistical assumptions underlying survey weights to adjust for sampling bias. In general, province estimates are likely to be the most reliable. In addition, low levels of most mutations and incomplete data for some demographic variables reduced the sample size available for evaluating associations between mutations and demographics. Allele frequencies were only estimated at the district level given that the panmixia assumption is unlikely to hold at larger scales. Technical limitations of the molecular assay included challenges in phasing SNPs from independent amplicons for haplotype construction, which could be mitigated by using long amplicon sequencing approaches.

Molecular approaches are increasingly recognized as essential surveillance tools for guiding programmatic decision-making in malaria control and elimination. Using multiplex amplicon sequencing and a geographically comprehensive sampling approach, this study identified concerning levels of *k*13 mutations associated with ART-R across multiple Zambian provinces and transmission strata. Although the impact of the mutations identified in this study on resistance and treatment efficacy is unclear, their high frequencies in certain health facilities and districts, coupled with consistent increases of the P441L mutation noted in other studies, suggest they may confer a selective advantage in a country predominantly using ACTs for treatment and interventions. Targeted therapeutic efficacy and ex vivo susceptibility studies, as well as longitudinal and geographically representative molecular surveillance, will be necessary to fully understand the implications of these mutations and monitor their spread. Decision-makers must also prioritize strategies to prevent the emergence and limit the spread of partner drug resistance and other ART-R genotypes, safeguarding treatment efficacy and progress toward elimination goals.

## Data Availability

All data will be made available upon final publication

## Acknowledgments

We thank the participants, nurses, field supervisors and data managers who made this study possible. We thank Nokwethemba Kubheka and Ongeziwe Taku for laboratory assistance, and Nicholas Hathaway for bioinformatics technical assistance. We also thank members of the Malaria Elimination Initiative, the EPPIcenter, and Johns Hopkins University’s IDDynamics group for fruitful technical discussions.

## Co-Author Contact Information

Andrés Aranda-Díaz. EPPIcenter Research Program, Department of Medicine, University of California, San Francisco, United States of America

Sydney Mwanza. Biomedical Sciences Department, National Health Research and Training Institute, Ndola, Zambia.

Takalani I. Makhanthisa. Laboratory for Antimalarial Resistance Monitoring and Malaria Operational Research (ARMMOR), Centre of Emerging Zoonotic and Parasitic Diseases, National Institute for Communicable Diseases; Johannesburg, South Africa

Sonja Lauterbach. Laboratory for Antimalarial Resistance Monitoring and Malaria Operational Research (ARMMOR), Centre of Emerging Zoonotic and Parasitic Diseases, National Institute for Communicable Diseases; Johannesburg, South Africa

Faith De Amaral. EPPIcenter Research Program, Department of Medicine, University of California, San Francisco, United States of America

Mukosha Chisenga. SADC Malaria Elimination Eight Secretariat, Windhoek, Namibia

Brighton Mangena. SADC Malaria Elimination Eight Secretariat, Windhoek, Namibia

Isobel Routledge. Malaria Elimination Initiative, Global Health Group, University of California, San Francisco, United States of America

Blaženka Letinić. Laboratory for Antimalarial Resistance Monitoring and Malaria Operational Research (ARMMOR), Centre of Emerging Zoonotic and Parasitic Diseases, National Institute for Communicable Diseases; Johannesburg, South Africa

Bertha Kasonde. Biomedical Sciences Department, National Health Research and Training Institute, Ndola, Zambia.

Gershom Chongwe, National Health Research and Training Institute, Ndola, Zambia

Mulenga C. Mwenda, PATH-Malaria Control and Elimination Partnership in Africa, Lusaka, Zambia.

John M. Miller. PATH-Malaria Control and Elimination Partnership in Africa, Lusaka, Zambia. E- mail

Tricia Hibwato National Malaria Elimination Centre, Ministry of Health, Lusaka, Zambia.

Chirwa Jacob, National Malaria Elimination Centre, Ministry of Health, Lusaka, Zambia.

Busiku Hamainza, National Malaria Elimination Centre, Ministry of Health, Lusaka, Zambia. E- mail

Stephen Bwalya, National Malaria Elimination Centre, Ministry of Health, Lusaka, Zambia. E- mail

Japhet Chiwaula, National Malaria Elimination Centre, Ministry of Health, Lusaka, Zambia. E- mail

Japhet Matoba, National Malaria Elimination Centre, Ministry of Health, Lusaka, Zambia.

Chadwick Sikaala. SADC Malaria Elimination Eight Secretariat, Windhoek, Namibia

John Chimumbwa. SADC Malaria Elimination Eight Secretariat, Windhoek, Namibia

Amy Wesolowski. Department of Epidemiology, Johns Hopkins University Bloomberg School of Public Health, Baltimore, Maryland, United States of America.

Jennifer L. Smith. Malaria Elimination Initiative, Global Health Group, University of California, San Francisco, United States of America

Jaishree Raman. Laboratory for Antimalarial Resistance Monitoring and Malaria Operational Research (ARMMOR), Centre of Emerging Zoonotic and Parasitic Diseases, National Institute for Communicable Diseases; Wits Research Institute for Malaria, University of Witwatersrand; UP Institute for Sustainable Malaria Control, University of Pretoria; Johannesburg, South Africa

Moonga Hawela, National Malaria Elimination Centre, Ministry of Health, Lusaka, Zambia. E- mail

