## Supplementary material for "*Plasmodium falciparum* genomic surveillance reveals a diversity of *kelch13* mutations in Zambia": Figure S

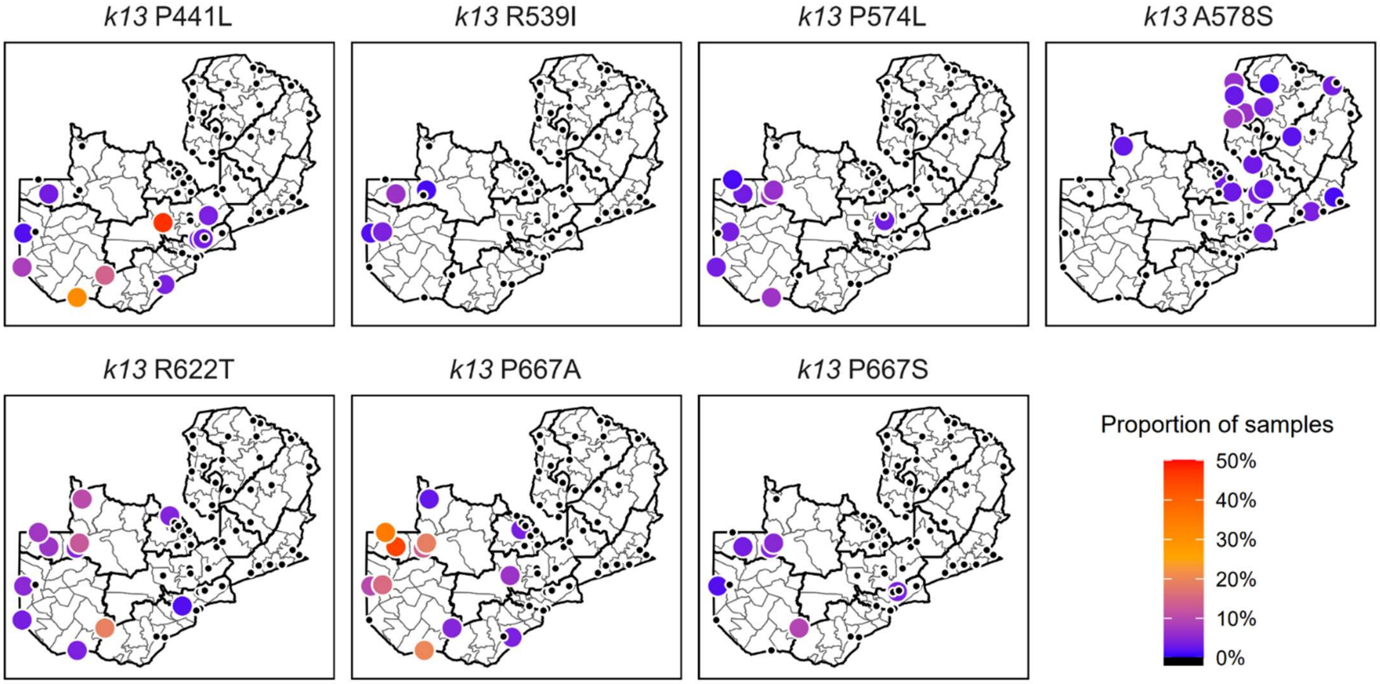


**Figure S1.** Proportion of samples carrying a *k13* mutation in each of the 61 health facilities.

Health facilities with non-zero proportions are shown in larger colored dots.

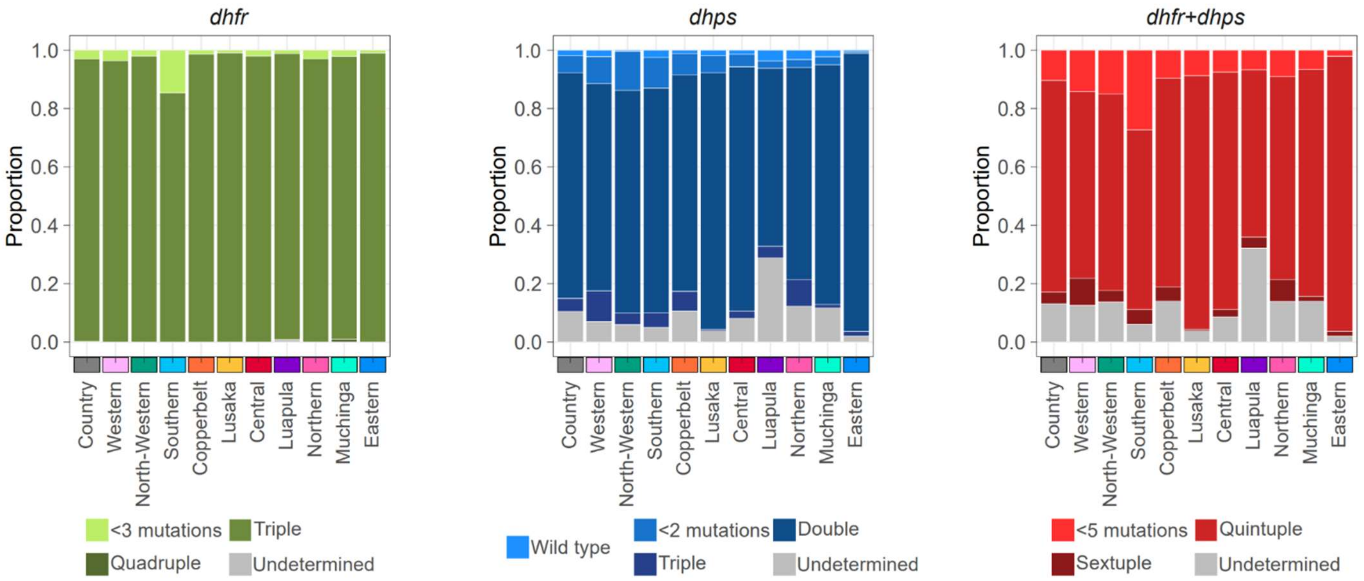


**Figure S2. Proportion of samples with *dhfr* and *dhps* haplotypes.** Proportion of samples carrying *dhfr*, *dhps*, or *dhfr*/*dhps* haplotypes. Each sample was assigned the haplotype corresponding to the highest level of resistance detected. Same as Figure 3C but including polyclonal samples with mixed genotypes in multiple targets classified as undetermined.
